# Development and validation of a deep learning algorithm based on fundus photographs for estimating the CAIDE dementia risk score

**DOI:** 10.1101/2021.08.17.21262156

**Authors:** Rong Hua, Jianhao Xiong, Gail Li, Yidan Zhu, Zongyuan Ge, Yanjun Ma, Meng Fu, Chenglong Li, Bin Wang, Li Dong, Xin Zhao, Zhiqiang Ma, Jili Chen, Chao He, Zhaohui Wang, Wenbin Wei, Fei Wang, Xiangyang Gao, Yuzhong Chen, Qiang Zeng, Wuxiang Xie

**Affiliations:** Peking University Clinical Research Institute, Peking University First Hospital, Beijing, China; PUCRI Heart and Vascular Health Research Center at Peking University Shougang Hospital, Beijing, China; Key Laboratory of Molecular Cardiovascular Sciences (Peking University), Ministry of Education, Beijing, China; Beijing Airdoc Technology Co., Ltd. Beijing, China; Departments of Psychiatry and Behavioral Sciences and Division of Gerontology and Geriatric Medicine, University of Washington, Seattle, WA, USA; Beijing Tongren Eye Center, Beijing Tongren Hospital, Beijing, China; iKang Guobin Healthcare Group Co., Ltd, Beijing, China; Shibei Hospital Jingan District, Shanghai, China; Health Management Institute, The Second Medical Center & National Clinical Research Center for Geriatric Diseases, Chinese PLA General Hospital

## Abstract

The Cardiovascular Risk Factors, Aging, and Incidence of Dementia (CAIDE) dementia risk score is a recognized tool for dementia risk stratification. However, its application is limited due to the requirements for multidimensional information and fasting blood draw. Consequently, effective, convenient and noninvasive tool for screening individuals with high dementia risk in large population-based settings is urgently needed. A deep learning algorithm based on fundus photographs for estimating the CAIDE dementia risk score was developed and internally validated by a medical check-up dataset included 271,864 participants in 19 province-level administrative regions of China, and externally validated based on an independent dataset included 20,690 check-up participants in Beijing. The performance for identifying individuals with high dementia risk (CAIDE dementia risk score ≥10 points) was evaluated by area under the receiver operating curve (AUC) with 95% confidence interval (CI). We found that the algorithm achieved an AUC of 0.944 (95% CI 0.939–0.950) in the internal validation group and 0.926 (95% CI, 0.913–0.939) in the external group, respectively. Besides, the estimated CAIDE dementia risk score derived from the algorithm was significantly associated with both comprehensive cognitive function and specific cognitive domains. In conclusion, this algorithm trained via fundus photographs could well identify individuals with high dementia risk in a population setting. Therefore, it has potential to be utilized as a noninvasive and more expedient method for dementia risk stratification. It might also be adopted in dementia clinical trials, incorporated as inclusion criteria to efficiently select eligible participants.

## Introduction

Worldwide, the number of people have dementia is projected to triply increase to 152 million by 2050, given the dramatic rise in ageing populations, yet there are no curative therapeutics available.^1^ Dementia has a long preclinical phase when no symptomatic cognitive impairments, but neurodegenerative progressions are occurring.^2^ Early identification of high-risk individuals is essential for preventing dementia, which efficiently targets participants who could benefit most from more intensive examinations and interventions.^3^

The Cardiovascular Risk Factors, Aging, and Incidence of Dementia (CAIDE) dementia risk score was a recognized model to predict 20-year dementia risk, which based on multidimensional risk factors: age, sex, educational level, physical inactivity, systolic blood pressure (SBP), total cholesterol (TC), and body mass index (BMI), with a total score ranged from 0 point to 15 points. It was also highly predictive in external validation of a large multiethnic population and adopted in Finnish Geriatric Intervention Study (FINGER) to select eligible at-risk participants. ^4-6^ However, the CAIDE dementia risk score entails measurements by questionnaire inquiry, physical examinations and fasting blood draw, these procedures are time-consuming or invasive for participants, also increase the labor costs of healthcare practitioners and produce biohazardous waste. Consequently, effective, convenient and noninvasive tool to screen individuals with high dementia risk in large population-based settings is warranted.

Vascular disease, especially microvasculature damage in the brain, is recognized as a major contributor to dementia.^1,7^ Anatomically and developmentally, the retina shares homology with the brain.^8^ The retina is an exceptional site where the microcirculation can be handily and noninvasively visualized by fundus photography, thus providing insights into the brain microvasculature. Large population studies have demonstrated the correlations between various retinal microvascular abnormalities (such as retinopathy, arteriolar narrowing and venular dilation) and increased risk of dementia.^9-11^ Moreover, the emerging artificial intelligence technique, especially deep learning, has realized integrating multiple retinal features from fundus photographs, to provide estimation on vascular risk factors, and prediction on cardiovascular diseases.^12,13^ However, to our knowledge, this method has not been investigated on predicting dementia.

Herein, we hypothesized that the deep learning algorithm trained via fundus photographs might help to dementia risk stratification. Due to the insufficient time length to occur enough dementia events in our dataset, the present study aimed to train a deep learning algorithm for estimating the CAIDE dementia risk score thus identifying individuals with high dementia risk, and we proposed that the estimated score generated from the algorithm associated with the cognitive function.

## Results

### Study population

The characteristics of individuals in the development dataset, internal validation dataset, and the external validation dataset were summarized in Table 1.

**Table 1.**
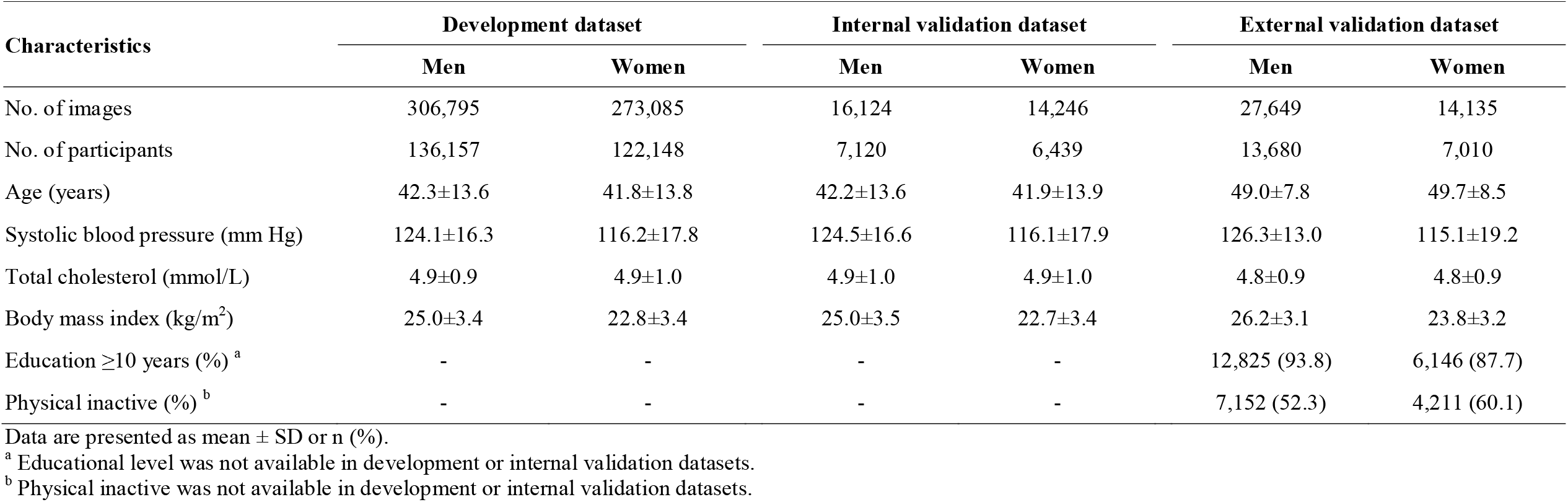
Characteristics of individuals in development, internal validation, and external validation datasets.

Among the 271,864 check-up participants, we randomly divided 95% (258,305 participants, mean aged 42.1 ± 13.4 years, men: 52.7%) into the development group and 5% (13,559 participants, mean aged 41.2 ± 13.3 years, men: 52.5%) into the internal validation group (eFigure 1a). These two groups shared similar baseline characteristics as shown in Table 1. Besides, the characteristics of participants in the training and tuning groups were displayed in eTable 1. Among 30,455 participants from the Chinese PLA General Hospital and its surrounding communities, 20,690 participants (mean aged 49.2 ± 8.0 years, men: 68.4%) had fundus photographs and complete information for calculating CAIDE dementia risk score and thus were included in the external validation group (eFigure 1b). Compared with the development population, individuals in the external validation were older.

Respectively, 200 (1.5%) individuals in the internal validation dataset and 236 (1.1%) in the external were in high dementia risk, with their CAIDE dementia risk score ≥10 points.

### Algorithm performance

The R^2^ between the estimated and actual CAIDE dementia risk score was 0.80 in the internal and 0.58 in the external validation datasets (Figure 1). As shown in Figure 2, the algorithm achieved an AUC of 0.944 (95% CI 0.939–0.950) in the internal validation dataset and 0.926 (95% CI, 0.913–0.939) in the external for identifying individuals with high dementia risk. The maximum Youden index on the two receiver operating characteristic curves were 0.801 with the sensitivity of 0.959 and specificity of 0.842, corresponded to the optimal cut-off point of 6.793 in the internal validation dataset, and 0.700 with the sensitivity of 0.911 and specificity of 0.789, corresponded to the optimal cut-off point of 6.714 in the external validation dataset, respectively.

**Figure 1.**
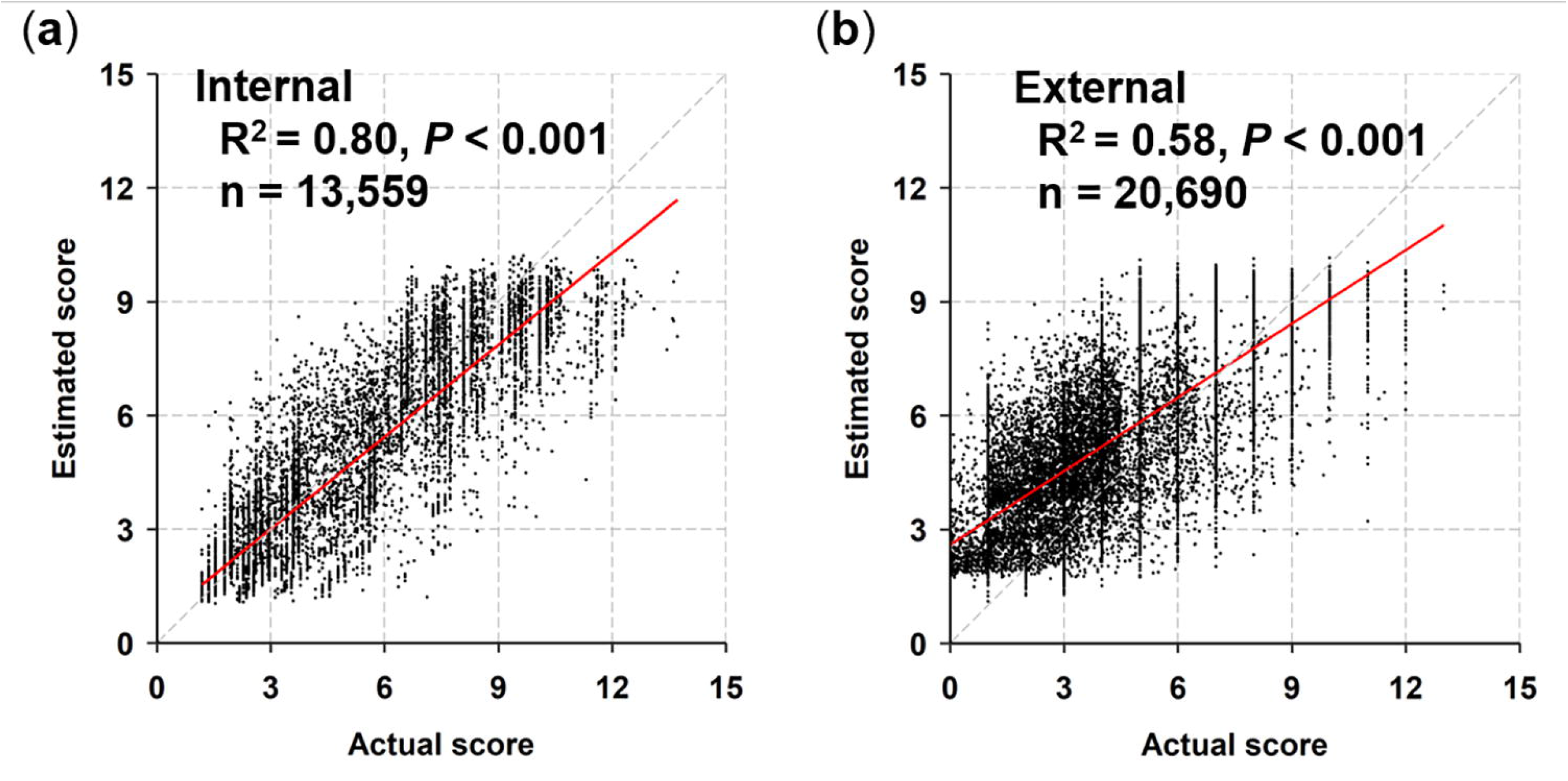
Estimation of CAIDE dementia risk score in the internal (a) and external (b) validations. The R^2^ (coefficient of determination) between the estimated CAIDE dementia risk score and actual CAIDE dementia risk score is presented.

**Figure 2.**
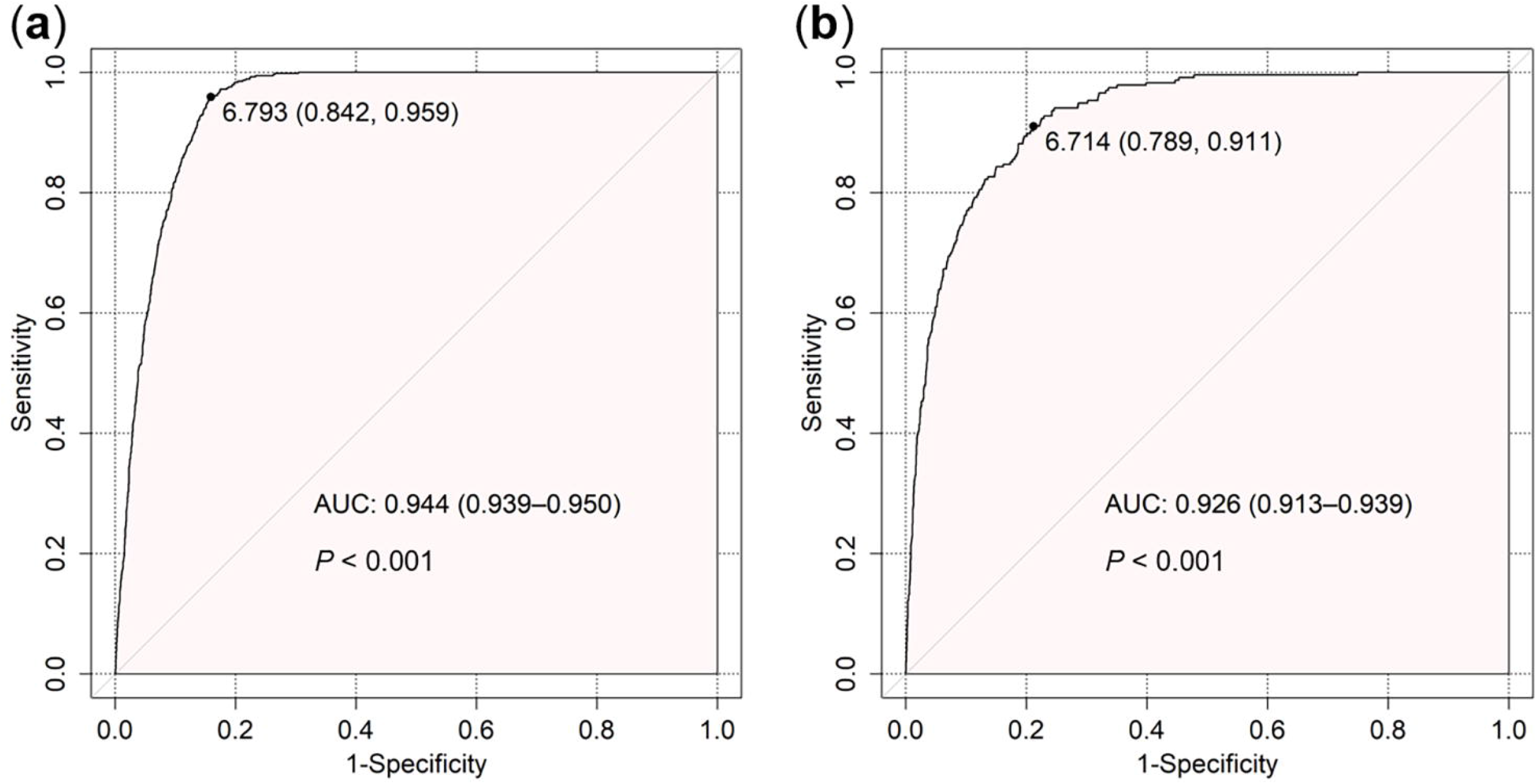
Algorithm performance for identifying participants with high dementia risk in the internal (a) and external (b) validations. Individuals with high dementia risk were defined as CAIDE dementia risk score ≥10 points. The points on line indicate the maximum Youden index. Abbreviation: AUC = area under the receiver operating characteristic curve.

### The estimated score and cognitive function

The characteristics of participants who received detailed cognitive assessments were presented in eTable 2. Linear regression analyses found that the estimated CAIDE dementia risk score (as continuous variable) was significantly associated with the score of MoCA. As shown in Table 2, 1-point increment of estimated CAIDE dementia risk score was significantly associated with −0.565 (95% CI, −0.673 to −0.457) increment of the MoCA score after multivariable adjustment, which manifested worse comprehensive cognitive performance. Similarly, the higher estimated CAIDE dementia risk score was significantly associated with lower score of memory and verbal fluency test, which indicated poorer performance of memory, language and executive function. The higher estimated score was also significantly associated with longer TMT-A and TMT-B time, which represented worse attention and executive function. The analysis of covariance found that after full adjustment, compared with the lowest quartile, the second, third, and highest quartiles were associated with worse comprehensive cognitive function, with lower MoCA score by −0.989 (95% CI, −1.452 to −0.525), −1.685 (95% CI, −2.158 to −1.212), and −2.247 (95% CI, −2.722 to −1.772), respectively (*P* for linear trend < 0.001, Table 3). Similar trends were also observed in performance of memory test, verbal fluency test, TMT-A and TMT-B.

**Table 2.**
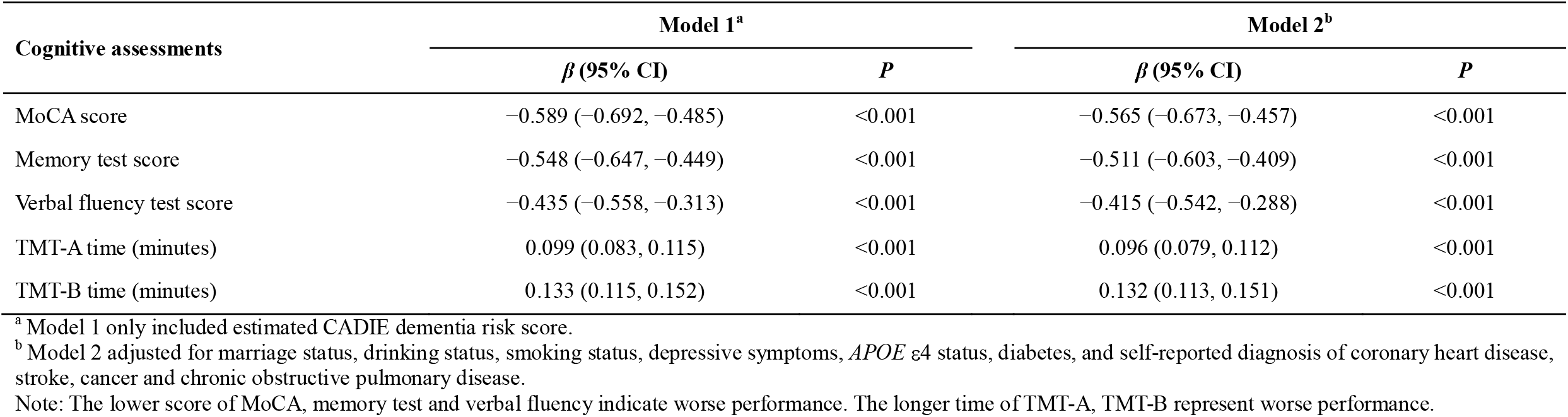
Association between estimated CAIDE dementia risk score and different cognitive assessments: using multiple linear regression models.

**Table 3.** Association between quartiles of estimated CAIDE dementia risk score and different cognitive assessments: using analysis of covariance.

| Estimated CAIDE dementia risk score | MoCA score |  | Memory test score |  | Verbal fluency test score |  | TMT-A time (minutes) |  | TMT-B time (minutes) |  |
| --- | --- | --- | --- | --- | --- | --- | --- | --- | --- | --- |
| | $\beta$ (95% CI) | *P* | $\beta$ (95% CI) | *P* | $\beta$ (95% CI) | *P* | $\beta$ (95% CI) | *P* | $\beta$ (95% CI) | *P* |
| Model 1<sup>a</sup> |  |  |  |  |  |  |  |  |  |  |
| Quartile 1 | 0.000 (ref.) | / | 0.000 (ref.) | / | 0.000 (ref.) | / | 0.000 (ref.) | / | 0.000 (ref.) | / |
| Quartile 2 | -1.053 (-1.512, -0.594) | <0.001 | -1.024 (-1.460, -0.587) | <0.001 | -0.561 (-1.101, -0.021) | <0.001 | 0.127 (0.057, 0.198) | <0.001 | 0.202 (0.121, 0.284) | <0.001 |
| Quartile 3 | -1.778 (-2.237, -1.319) | <0.001 | -1.500 (-1.937, -0.063) | <0.001 | -1.249 (-1.789, -0.804) | <0.001 | 0.253 (0.182, 0.323) | <0.001 | 0.367 (0.285, 0.449) | <0.001 |
| Quartile 4 | -2.354 (-2.814, -1.896) | <0.001 | -2.220 (-2.656, -1.783) | <0.001 | -1.630 (-2.170, -1.089) | <0.001 | 0.391 (0.320, 0.461) | <0.001 | 0.510 (0.428, 0.592) | <0.001 |
| Test for linear trend | -0.779 (-0.924, -0.634) | <0.001 | -0.713 (-0.852, -0.575) | <0.001 | -0.558 (-0.728, -0.387) | <0.001 | 0.130 (0.107, 0.152) | <0.001 | 0.169 (0.144, 0.195) | <0.001 |
| Model 2<sup>b</sup> |  |  |  |  |  |  |  |  |  |  |
| Quartile 1 | 0.000 (ref.) | / | 0.000 (ref.) | / | 0.000 (ref.) | / | 0.000 (ref.) | / | 0.000 (ref.) | / |
| Quartile 2 | -0.989 (-1.452, -0.525) | <0.001 | -0.917 (-1.356, -0.478) | <0.001 | -0.532 (-1.075, 0.011) | 0.055 | 0.122 (0.052, 0.193) | <0.001 | 0.208 (0.126, 0.290) | <0.001 |
| Quartile 3 | -1.685 (-2.158, -1.212) | <0.001 | -1.343 (-1.791, -0.895) | <0.001 | -1.199 (-1.754, -0.645) | <0.001 | 0.243 (0.171, 0.316) | <0.001 | 0.370 (0.287, 0.454) | <0.001 |
| Quartile 4 | -2.247 (-2.722, -1.772) | <0.001 | -2.065 (-2.515, -1.615) | <0.001 | -1.538 (-2.094, -0.981) | <0.001 | 0.376 (0.303, 0.449) | <0.001 | 0.502 (0.418, 0.586) | <0.001 |
| Test for linear trend | -0.742 (-0.893, -0.591) | <0.001 | -0.662 (-0.805, -0.519) | <0.001 | -0.527 (-0.704, -0.350) | <0.001 | 0.125 (0.102, 0.148) | <0.001 | 0.167 (0.140, 0.193) | <0.001 |

<sup>a</sup> Model 1 only included estimated CADIE dementia risk score.
<sup>b</sup> Model 2 adjusted for marriage status, drinking status, smoking status, depressive symptoms, *APOE* $\epsilon 4$ status, diabetes, and self-reported diagnosis of coronary heart disease, stroke, cancer and chronic obstructive pulmonary disease.
Note: The lower score of MoCA, memory test and verbal fluency indicate worse performance. The longer time of TMT-A, TMT-B represent worse performance.

### Sensitivity analysis

As shown in eFigure 2, the algorithm still performed well in screening individuals with high dementia risk when the cut-off score changed to 9 points, with an AUC of 0.947 (95% CI, 0.942–0.951) in the internal validation dataset and 0.896 (95% CI, 0.886–0.906) in the external validation. As shown in eFigure 3, the algorithm exhibited moderate performance in identifying participants eligible for multidomain intervention, with an AUC of 0.977 (95% CI, 0.975–0.980) in the internal validation dataset and 0.857 (95% CI, 0.851–0.863) in the external validation. Besides, eFigure 4 summarized the algorithm performance in subgroups of the external validation. The algorithm presented a higher AUC in female (0.808 vs 0.733, *P* = 0.049), as well as in participants <60 years (0.806 vs 0.703, *P* = 0.009). As eFigure 5 presented, we found no interaction effect of sex or age group on the associations between estimated CAIDE dementia risk score and the score of MoCA, or other specific cognitive functions.

## Discussion

To the best of our knowledge, the present study is the first investigation on developing a deep learning algorithm based on fundus photographs for identifying individuals with high dementia risk, with an AUC of 0.944 (95% CI 0.939–0.950) in the internal validation, and 0.926 (95% CI, 0.913–0.939) in the external validation dataset.

Moreover, the estimated CAIDE dementia risk score exhibited significant associations with both comprehensive and specific domains of cognitive function, which further supported the reasonability of the algorithm. Taken together, our study clarified the feasibility of adopting deep learning algorithm based on fundus photographs to screen individuals with high dementia risk in population-based settings.

The rationale of our work based on the concept that, the retina shares similar morphological features and physiological properties with the brain, and hence provide a unique site to detect changes in microvasculature related to the development of dementia.^8^ Previous studies have investigated the associations between a spectrum of retinal vascular abnormalities measured via fundus photography and the risk of dementia.^9-11^ However, most studies measured retinal signs by semi-automated software, requiring human identification on the basis of prespecified protocols, which might introduce intra- and inter-variability. Besides, recent systematic reviews indicated that combination of multiple retinal vascular parameters, rather than individual marker, might provide higher prognostic value.^14,15^ The present study utilized artificial intelligence technique, which might exhibit notable advantages in these issues. Artificial intelligence operates in absence of human assessment, and even performs superiorly to ophthalmologists in capturing subtle retinal changes that would otherwise fail to attract human attention.^16^ With faster, easier, more consistent and precise output, the artificial intelligence reduces variability and human cost, thus enhancing the clinical utility of retinal photography.^17^ Moreover, artificial intelligence is able to fully extract and integrate multiple retinal features (including information beyond human existed perception or understanding) that are related to dementia risk.

Participants in the external validation were older, and had a larger proportion of male. The demographic heterogeneity between the development dataset and the external validation dataset suggested the algorithm’s robustness and promising wider utility. One application scenario for the algorithm is screening individuals with high dementia risk in community. Traditional dementia prediction models requiring cognitive tests or multidimensional risk factors increased application difficulties in population-based settings. By contrast, fundus photography is easy to implement and timesaving. According to our practical experience, an investigator with no background on ophthalmology could take fundus photographs within one minute after a few hours of training. Besides, compared with risk factors like blood lipids or glucose, the retinal images have no requirement for fasting status, with less fluctuation and can be obtained noninvasively, thus facilitating the acceptability and convenience of participants. In addition, the algorithm could also be recommended as an add-on to routine screening for diabetic retinopathy, given that patients with diabetes were significantly associated with higher risk of cognitive decline and dementia.^18^ Moreover, our algorithm has potential utility in assessing pre-test dementia probability for further diagnostic tests in outpatient clinics. Last but not the least, this algorithm might also be adopted in dementia clinical trials, incorporated as inclusion criteria to efficiently target eligible participants, or surrogate outcome which could be observed expediently.^19^

Previous studies have investigated deep learning algorithm based on fundus photographs for screening cardiovascular diseases and anaemia,^12,13,20^ our study added novel evidence regarding dementia in this field, potentially facilitating the eventual application of fundus photography for simultaneous screening of multiple diseases in large population-based settings. The foremost strength of the present work was employing convolutional neural network to deal with large dataset of fundus images. The development dataset contained 579,880 fundus images of 258,305 individuals from 19 province-level administrative regions of China, the convolutional neural network exhibited distinct advantages in processing such large dataset, by extracting information from images with a deep architecture, which was similar to image process in human brain.^21^ Another strength was incorporating external validation cohort with varied demographic characteristics and comprehensive cognitive tests, the results externally validated the performance and further supported the scientificalness of the algorithm.

There were, however, also limitations in our study. First, the CAIDE dementia risk score was derived from cross-sectional data, investigations based on incident dementia events in longitudinal settings are warranted to further verify the predictive ability of the algorithm. Second, the R^2^ in the external validation was relatively lower, probably due to the distinct age difference between the development and external validation datasets, given that age is the most important factor for dementia and cognitive function. Another reason could be the absence of educational level and physical inactivity in the development dataset, which was conducted in medical check-up settings mainly collecting health measurements rather than socioeconomic factors or lifestyle. Future collection of information about educational level and physical inactivity in the development dataset could improve the algorithm’s performance. Third, the present study only included Chinese participants, which might limit the generalization of our algorithm to other ethnicities.

## Conclusions

The present study demonstrated that a deep learning algorithm based on fundus photographs could well identify individuals with high dementia risk, and hold promise for wider application in community-based screening or clinic. As far as we know, this work is the first attempt to utilize deep learning technology and fundus photographs for screening dementia, future advancements in artificial intelligence technology and larger collection of relevant data would further improve and verify the performance of the algorithm.

## Materials and methods

### Study design

This was a cross-sectional study. A deep learning algorithm based on fundus photographs for estimating the CAIDE dementia risk score was developed and internally validated by a medical check-up dataset. Additionally, by an independent dataset, we externally validated the algorithm’s discrimination on individuals with high dementia risk, and further explored the association between the estimated CAIDE dementia risk score and cognitive function.

### Participants and datasets

For the algorithm development, a dataset from 271,864 participants from Tongren Hospital in Beijing, Shibei Hospital in Shanghai, and iKang Healthcare Group who attending medical check-up in 19 province-level administrative regions of China during September 2018 to December 2019, were randomly divided into development (95%) and internal validation (5%) components. This dataset contained retinal fundus images and routine medical information, including age, sex, SBP, TC, and BMI. The use of the dataset for the algorithm training was approved by Tongren Hospital Institutional Review Board, Shibei Hospital Institutional Review Board, and iKang Healthcare Group Institutional Review Board with a waiver of informed consent. The algorithm’s performance was further externally validated using data derived from adults attending the medical check-up in Chinese PLA General Hospital and surrounding communities during October 2009 to December 2020. The use of this dataset was approved by Chinese PLA General Hospital Institutional Review Board (ethical review approval number: S2019-131-01) and Peking University School Institutional Review Board (ethical review approval number: IRB0001052–19060), all participants have given written informed consent.

Both datasets collected fundus images and clinical characteristics on the same day.

A variety of digital nonmydriatic fundus cameras were adopted to obtain fundus images, including Canon CR1/CR2 and Crystalvue FundusVue/ TonoVue in the development dataset, Canon CR1 and Centervue DRS in the external validation dataset. All images were captured using 45° fields of view. Both datasets calculated the CAIDE dementia risk score based on the function proposed by Kivipelto et al.^4^

However, educational level and physical inactivity were not collected in the check-up dataset. We imputed the risk score of educational level to the algorithm based on the Sixth National Census,^22^ according to the average risk score of educational level among the corresponding sex and age group of the individual. Score of physical inactivity was imputed according to BMI status, those with a BMI ≥24 kg/m^2^ were regarded as physical inactive, given that higher BMI was significantly associated with physical inactivity.^23^

### Development of the algorithm

The development dataset consists of a training dataset and a tuning dataset. The training dataset was used to update model parameters during the training stage, and the tuning set was used for model selection. The label for training and testing of the network is given as y_CAIDE Score_ which is the score summation of risk factors according to the CAIDE dementia risk model.^4^

Our CAIDE algorithm was trained and tested using InceptionResNetV2 architecture on the platform Keras v2.2.2 and the Python scikit-learn package 0.22.2. The open source frameworks platform Keras v2.2.2 was available at https://github.com/keras-team/keras. The source code of InceptionResNetV2 was obtained from https://github.com/keras-team/keras-applications/blob/master/keras_applications/inception_resnet_v2.py. The training and testing of the algorithm were performed using a GTX 1080Ti GPU ×2 (CUDA version 9.0, Nvidia Corp., USA) with a batch size of 64 on an operation system Ubuntu v16.04.6. The model was trained for prediction of the CAIDE score as a regression task. We deployed Mean Absolute Error (MAE) as the loss function to minimize during the training stage by Adam optimizer.^24^

The image data was loaded by using OpenCV version 4.2.0. The data augmentations of random cropping, random rotation (±30°) and random horizontal flipping were implemented by Keras image augmentation package of data generator. In order to improve the robustness of model performance on varying image quality and photography style. An image normalization method, enhanced domain transformation, was used to map any input image pixel values to a given task distribution.^25^ To speed up training and validation, multi-processing and 12 workers were utilized by implementing Keras fit generator function.

### Validation of the algorithm

The estimated CAIDE dementia risk score of the participants deprived from mean estimated y_CAIDE Score_ of both eyes, and the actual dementia risk score was calculated according to the CAIDE model. The goodness of fit of the algorithm was assessed by adjusted coefficient of determination (R^2^) in the internal and external validation datasets. Besides, the algorithm’s discrimination on identifying individuals with high dementia risk was evaluated by area under the receiver operating curve (AUC) with 95% confidence interval (CI) by the pROC package version 1.16.2. Consistent with Sindi et al, dementia risk score ≥10 points was recognized as high dementia risk.^6^ The maximum Youden index was applied to determine the optimal cut-off point.

### Cognitive assessments

Among the external validation population, 1,512 middle-aged and older participants have received detailed cognitive tests. We further explored the associations between the estimated CAIDE dementia risk score and cognitive function based on this population. The primary cognitive measurement was the Chinese version of Montreal Cognitive Assessment (MoCA) Basic, a sensitive and validated cognitive test battery to comprehensively assess nine cognitive domains.^26^ In addition, we also supplemented three tests to further assess specific cognitive domains. Specifically, the memory function was measured by immediate and delayed recall of a list of ten unrelated words, and the total score ranged from 0 to 20.^27^ The language and executive function was assessed by a verbal fluency test, which requiring participants to speak names of animals as many as possible within 1 minute, and the total number of animal names (excluding repetitive names) was count as the test score.^28^ The attention function and executive function were evaluated by the Chinese version of Trails Making Test (TMT),^29^ which asking individuals to draw a line through 25 numbers consecutively in ascending order, and as fast as they could. The TMT included two tasks, the TMT-A comprised numbers from 1 to 25, while the TMT-B was different in 25 numbers enclosed in squares from 1 to 12 and circles from 1 to 13. The TMT-A evaluated processing speed and visual attention, and the TMT-B assessed executive function by measuring cognitive alternation ability. In both tests of memory and verbal fluency, the higher score indicated better cognitive performance, while in the TMT, the longer time manifested worse performance.

### Statistical analysis

The results were presented using percentage for categorical variables and means ± standard deviations (SD) for continuous variables. We ran multiple linear regression models to examine the associations between the estimated CAIDE dementia risk score and different cognitive assessments. The first model only included the estimated score, while the second model adjusted for multiple covariates, which contained marriage status, drinking status, smoking status, depressive symptoms, *APOE* ε4 status, and chronic diseases status. Specifically, marriage status indicated currently married or not. Participants were divided into non-smokers (including ex-smokers) and current smokers. Alcohol consuming was defined as drinking at least once per week over the past one year. We employed the ten-item version of the Center for Epidemiologic Studies Depression Scale (CES-D) to assess depressive symptoms, with a summed score ranged from 0 to 30. According to the prior study, a score ≥12 was defined as having depressive symptoms in our study.^30^ Individuals were divided into *APOE* ε4 carriers (indicated the presence of one or two ε4 alleles) and noncarriers. Diabetes was defined as HbA_1c_ ≥6.5% or fasting blood glucose ≥7.0 mmol/L, or self-reported current use of anti-diabetic therapy. Chronic disease measures also included self-reported physician-diagnosed coronary heart disease, cancer, stroke, and chronic obstructive pulmonary disease. Besides, we also employed analysis of covariance to compare cognitive performance between quartiles of the estimated dementia risk score, with the lowest quartile as the reference. Linear trend was also tested by including risk score quartiles as numerical variables.

To test the robustness of the algorithm, we evaluated the performance of the algorithm using 9 points as the cut-off score of high dementia risk, in consistent with a previous study.^31^ We further tested the ability of the algorithm to identify participants eligible for multidomain intervention, since the FINGER trial adopted CAIDE score ≥6 points as one of the inclusion criteria to select eligible at-risk participants among the general population.^19^ In addition, we conducted subgroup analyses according to sex, age group (<60 years and ≥60 years), respectively, based on the external validation dataset. For algorithm performance in identifying high risk individuals (with CAIDE score ≥10 points), we used Delong test to compare the AUC between subgroups. For the association with cognitive function, we respectively included the interaction terms of estimated dementia risk score with sex, as well as age group in multivariate linear regression models.

All statistical analyses were performed by SAS 9.4 (SAS Institute, Cary, NC), and R language 4.0.0 (R Foundation, Vienna, Austria), with two-tailed alpha value of 0.05 as the statistically significant level.

## Supporting information

eTable eFigure

## Data Availability

Individual participant data will be made available upon reasonable request, directed to the corresponding author (WX and QZ). Data can be shared through a secure online platform for research purposes. We applied the open-source machine-learning framework InceptionResNetV2 to do the experiments. Considering that many aspects of the experimental system (like data generation and model training) largely depend on our internal infrastructure, tooling, and hardware, we are unable to publicly release the code in the present stage. However, the experiments and implementation approaches are provided in the methods section.

## Acknowledgments

We thank all participants in the development dataset and external validation datasets. The present work was supported by National Natural Science Foundation of China (project no. 81974489), 2019 Irma and Paul Milstein Program for Senior Health Research Project Award, National Key R&D Programme of China (2017YFE0118800).

## Description of supplemental information

The supplemental material included 2 tables and 5 figures.

## Reference

1. Livingston, G., Huntley, J., Sommerlad, A., et al. (2020). Dementia prevention, intervention, and care: 2020 report of the Lancet Commission. Lancet 396, 413–446, 10.1016/S0140-6736(20)30367-6.

2. Sperling, R.A., Aisen, P.S., Beckett, L.A., et al. (2011). Toward defining the preclinical stages of Alzheimer’s disease: recommendations from the National Institute on Aging-Alzheimer’s Association workgroups on diagnostic guidelines for Alzheimer’s disease. Alzheimers Dement 7, 280–292, 10.1016/j.jalz.2011.03.003.

3. Kivipelto, M., Mangialasche, F., and Ngandu, T. (2018). Lifestyle interventions to prevent cognitive impairment, dementia and Alzheimer disease. Nature Reviews Neurology 14, 653–666, 10.1038/s41582-018-0070-3.

4. Kivipelto, M., Ngandu, T., Laatikainen, T., et al. (2006). Risk score for the prediction of dementia risk in 20 years among middle aged people: a longitudinal, population-based study. Lancet Neurol 5, 735–741, 10.1016/S1474-4422(06)70537-3.

5. Exalto, L.G., Quesenberry, C.P., Barnes, D., et al. (2014). Midlife risk score for the prediction of dementia four decades later. Alzheimers Dement 10, 562–570, 10.1016/j.jalz.2013.05.1772.

6. Sindi, S., Calov, E., Fokkens, J., et al. (2015). The CAIDE Dementia Risk Score App: The development of an evidence-based mobile application to predict the risk of dementia. Alzheimers Dement (Amst) 1, 328–333, 10.1016/j.dadm.2015.06.005.

7. Cheung, C.Y.L., Ikram, M.K., Chen, C., and Wong, T.Y. (2017). Imaging retina to study dementia and stroke. Prog Retin Eye Res 57, 89–107, 10.1016/j.preteyeres.2017.01.001.

8. Patton, N., Aslam, T., Macgillivray, T., et al. (2005). Retinal vascular image analysis as a potential screening tool for cerebrovascular disease: a rationale based on homology between cerebral and retinal microvasculatures. J Anat 206, 319–348, 10.1111/j.1469-7580.2005.00395.x.

9. Lesage, S.R., Mosley, T.H., Wong, T.Y., et al. (2009). Retinal microvascular abnormalities and cognitive decline The ARIC 14-year follow-up study. Neurology 73, 862–868, 10.1212/WNL.0b013e3181b78436.

10. de Jong, F.J., Schrijvers, E.M., Ikram, M.K., et al. (2011). Retinal vascular caliber and risk of dementia: the Rotterdam study. Neurology 76, 816–821, 10.1212/WNL.0b013e31820e7baa.

11. Deal, J.A., Sharrett, A.R., Albert, M., et al. (2019). Retinal signs and risk of incident dementia in the Atherosclerosis Risk in Communities study. Alzheimers Dement 15, 477–486, 10.1016/j.jalz.2018.10.002.

12. Poplin, R., Varadarajan, A.V., Blumer, K., et al. (2018). Prediction of cardiovascular risk factors from retinal fundus photographs via deep learning. Nature Biomedical Engineering 2, 158–164, 10.1038/s41551-018-0195-0.

13. Cheung, C.Y., Xu, D., Cheng, C.-Y., et al. (2021). A deep-learning system for the assessment of cardiovascular disease risk via the measurement of retinal-vessel calibre. Nature Biomedical Engineering 5, 498–508, 10.1038/s41551-020-00626-4.

14. McGrory, S., Cameron, J.R., Pellegrini, E., et al. (2016). The application of retinal fundus camera imaging in dementia: A systematic review. Alzheimers Dement (Amst) 6, 91–107, 10.1016/j.dadm.2016.11.001.

15. Wagner, S.K., Fu, D.J., Faes, L., et al. (2020). Insights into Systemic Disease through Retinal Imaging-Based Oculomics. Transl Vis Sci Technol 9, 6–6, 10.1167/tvst.9.2.6.

16. Son, J., Shin, J.Y., Kim, H.D., et al. (2020). Development and Validation of Deep Learning Models for Screening Multiple Abnormal Findings in Retinal Fundus Images. Ophthalmology 127, 85–94, 10.1016/j.ophtha.2019.05.029.

17. Ting, D.S.W., Cheung, C.Y.L., Lim, G., et al. (2017). Development and Validation of a Deep Learning System for Diabetic Retinopathy and Related Eye Diseases Using Retinal Images From Multiethnic Populations With Diabetes. Jama-J Am Med Assoc 318, 2211–2223, 10.1001/jama.2017.18152.

18. Zheng, F.F., Yan, L., Yang, Z.C., et al. (2018). HbA(1c), diabetes and cognitive decline: the English Longitudinal Study of Ageing. Diabetologia 61, 839–848, 10.1007/s00125-017-4541-7.

19. Ngandu, T., Lehtisalo, J., Solomon, A., et al. (2015). A 2 year multidomain intervention of diet, exercise, cognitive training, and vascular risk monitoring versus control to prevent cognitive decline in at-risk elderly people (FINGER): a randomised controlled trial. Lancet 385, 2255–2263, 10.1016/S0140-6736(15)60461-5.

20. Mitani, A., Huang, A., Venugopalan, S., et al. (2020). Detection of anaemia from retinal fundus images via deep learning. Nature Biomedical Engineering 4, 18–27, 10.1038/s41551-019-0487-z.

21. Anwar, S.M., Majid, M., Qayyum, A., et al. (2018). Medical Image Analysis using Convolutional Neural Networks: A Review. J Med Syst 42, 226, 10.1007/s10916-018-1088-1.

22. Tubulation on the 2010 population census of the People’s Republic of China (2010). http://www.stats.gov.cn/english/Statisticaldata/CensusData/rkpc2010/indexch.htm.

23. Tian, Y., Jiang, C., Wang, M., et al. BMI, leisure-time physical activity, and physical fitness in adults in China: results from a series of national surveys, 2000–14.

24. DeHoog, E., and Schwiegerling, J. (2008). Optimal parameters for retinal illumination and imaging in fundus cameras. Appl Optics 47, 6769–6777, Doi 10.1364/Ao.47.006769.

25. Xiong, J.H., He, A.W., Fu, M., et al. (2020). Improve Unseen Domain Generalization via Enhanced Local Color Transformation. International Conference on Medical Image Computing and Computer-Assisted Intervention.

26. Huang, L., Chen, K.L., Lin, B.Y., et al. (2018). Chinese version of Montreal Cognitive Assessment Basic for discrimination among different severities of Alzheimer’s disease. Neuropsych Dis Treat 14, 2133–2140, 10.2147/Ndt.S174293.

27. Hua, R., Ma, Y.J., Li, C.L., et al. (2021). Low levels of low-density lipoprotein cholesterol and cognitive decline. Science Bulletin 16, 1684–1690, https://doi.org/10.1016/j.scib.2021.02.018.

28. Xie, W.X., Zheng, F.F., Yan, L., and Zhong, B.L. (2019). Cognitive Decline Before and After Incident Coronary Events. Journal of the American College of Cardiology 73, 3041–3050, 10.1016/j.jacc.2019.04.019.

29. Wei, M.Q., Shi, J., Li, T., et al. (2018). Diagnostic Accuracy of the Chinese Version of the Trail-Making Test for Screening Cognitive Impairment. J Am Geriatr Soc 66, 92–99, 10.1111/jgs.15135.

30. Ma, Y., Liang, L., Zheng, F., et al. (2020). Association Between Sleep Duration and Cognitive Decline. JAMA Netw Open 3, e2013573–e2013573, 10.1001/jamanetworkopen.2020.13573.

31. Kaffashian, S., Dugravot, A., Elbaz, A., et al. (2013). Predicting cognitive decline: a dementia risk score vs. the Framingham vascular risk scores. Neurology 80, 1300–1306, 10.1212/WNL.0b013e31828ab370.

