## Supplementary material for "Development and validation of a deep learning algorithm based on fundus photographs for estimating the CAIDE dementia risk score": eTable eFigure

**eTable 1.** Characteristics of individuals in training and tuning datasets.

**eTable 2.** Characteristics of individuals received cognitive assessments.

**eFigure 1.** Flow chart of participants selection in development and internal validation datasets (a) and the external validation dataset (b).

**eFigure 2.** Algorithm performance for identifying individuals with CAIDE dementia risk score ≥9 points in the internal validation dataset (a) and the external validation dataset (b).

**eFigure 3.** Algorithm performance for identifying individuals with CAIDE dementia risk score ≥6 points in the internal validation dataset (a) and the external validation dataset (b).

**eFigure 4.** Algorithm performance for identifying participants with high dementia risk, subgroup results of sex (a) and age group (b) in the external validation.

**eFigure 5.** Association between estimated CAIDE dementia risk score and different cognitive assessments using multiple linear regression models, subgroup results of sex and age group in the external validation.

**eTable 1. Characteristics of individuals in training and tuning datasets**

| **Characteristics** | **Training dataset** | |  | **Tuning dataset** | |
| --- | --- | --- | --- | --- | --- |
|  | **Men** | **Women** |  | **Men** | **Women** |
| No. of images | 290,249 | 258,915 |  | 16,546 | 14,170 |
| No. of participants | 128,880 | 115,773 |  | 7,277 | 6,375 |
| Age (years) | 42.3±13.6 | 41.8±13.8 |  | 42.4±13.6 | 41.8±13.7 |
| Systolic blood pressure (mm Hg) | 124.0±16.3 | 116.2±17.9 |  | 124.7±16.3 | 116.3±17.6 |
| Total cholesterol (mmol/L) | 4.9±0.9 | 4.9±1.0 |  | 4.9±0.9 | 4.9±1.0 |
| Body mass index (kg/m^2^) | 25.0±3.4 | 22.8±3.4 |  | 24.9±3.4 | 22.8±3.4 |

Data are presented as mean ± SD or n (%).

Educational level and physical inactive were not available in training or tuning datasets.

**eTable 2. Characteristics of individuals received cognitive assessments**

| **Characteristics** | **Men** | **Women** |
| --- | --- | --- |
| No. of images | 1,122 | 1,902 |
| No. of participants | 561 | 951 |
| Age (years) | 60.7±7.1 | 59.3±7.3 |
| Systolic blood pressure (mm Hg) | 136.1±16.1 | 130.3±17.4 |
| Total cholesterol (mmol/L) | 4.8±0.9 | 5.0±0.9 |
| Body mass index (kg/m^2^) | 26.2±3.2 | 25.8±3.5 |
| Education ≥10 years (%) | 328 (58.5) | 576 (60.6) |
| Physical inactive (%) | 500 (89.1) | 739 (77.7) |
| Current smoking (%) | 299 (53.3) | 31 (3.3) |
| Current drinking (%) | 283 (50.4) | 46 (4.8) |
| Married (%) | 526 (93.8) | 818 (86.0) |
| Depressive symptoms (%) | 26 (4.6) | 68 (7.2) |
| *APOE* ε4 carrier (%) | 97 (17.3) | 165 (17.4) |
| History of diseases |  |  |
| Diabetes (%) | 175 (31.2) | 244 (25.7) |
| Coronary heart disease (%) | 51 (9.1) | 80 (8.4) |
| Stroke (%) | 47 (8.4) | 55 (5.8) |
| Cancer (%) | 18 (3.2) | 41 (4.3) |
| Chronic obstructive pulmonary disease (%) | 13 (2.3) | 7 (0.7) |
| Cognitive performance |  |  |
| MoCA score (point) | 24.0±3.3 | 25.0±3.3 |
| Memory test score (point) | 7.9±3.1 | 8.8±3.1 |
| Verbal fluency score (point) | 15.3±3.8 | 15.2±3.8 |
| TMT-A time (minute) | 1.2±0.5 | 1.1±0.5 |
| TMT-B time (minute) | 2.2±0.6 | 2.2±0.6 |

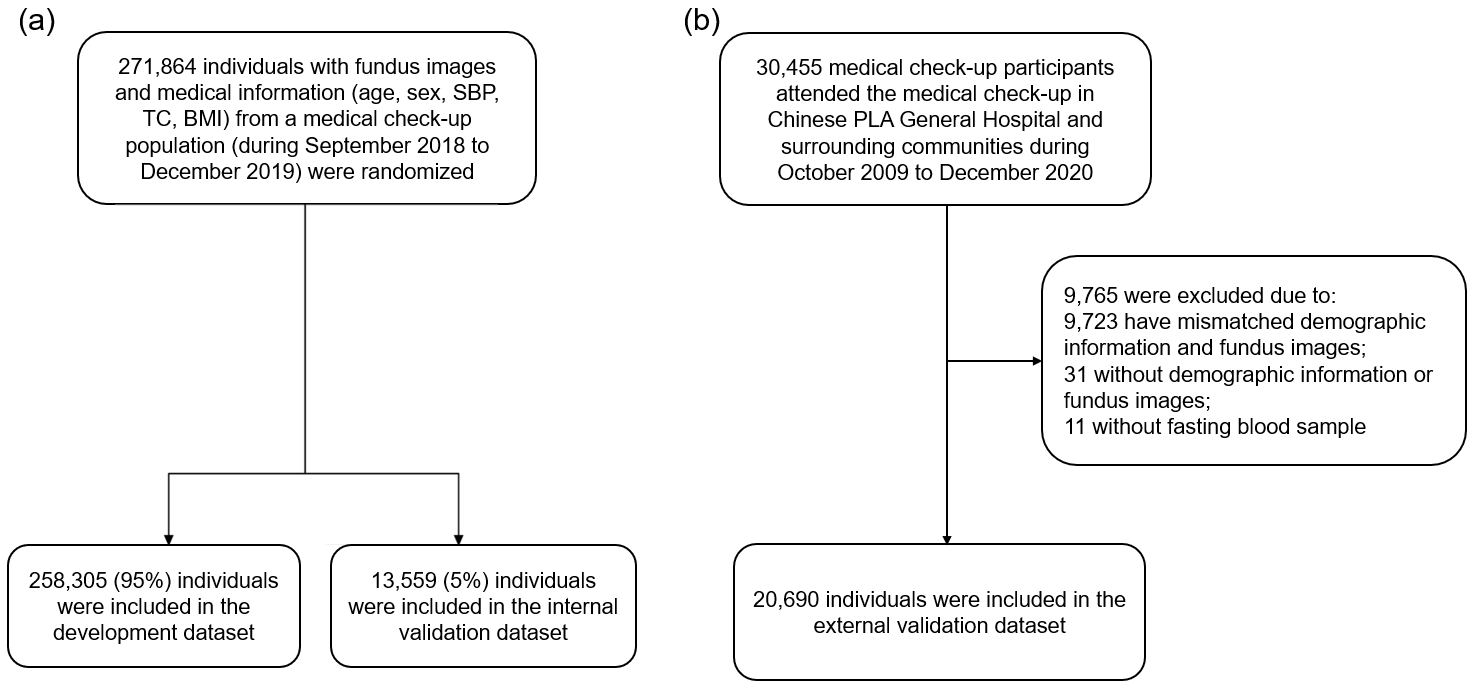

**eFigure 1. Flow chart of participants selection in development and internal validation datasets (a) and the external validation dataset (b).**

Abbreviations: SBP = systolic blood pressure. TC = total cholesterol. BMI = body mass index.

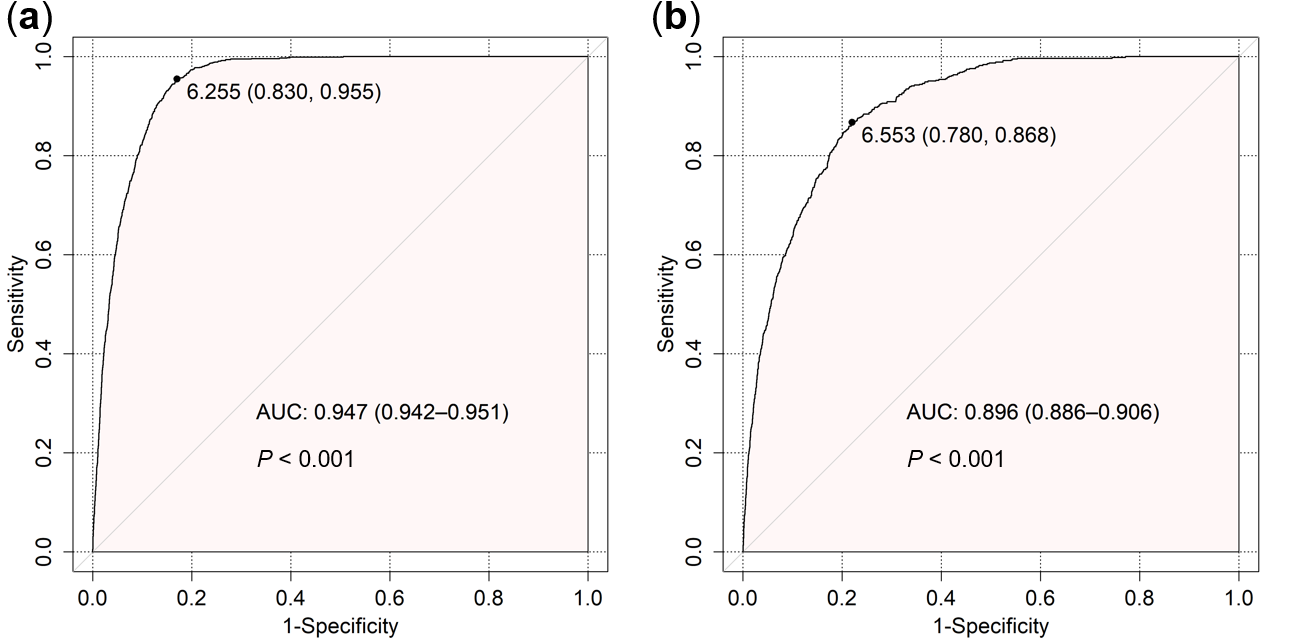

**eFigure 2. Algorithm performance for identifying individuals with CAIDE dementia risk score ≥9 points in the internal validation dataset (a) and the external validation dataset (b).**

Individuals with CAIDE dementia risk score ≥9 points were considered as having high dementia risk. The points on line indicate the maximum Youden index.

Abbreviation: AUC = area under the receiver operating characteristic curve.

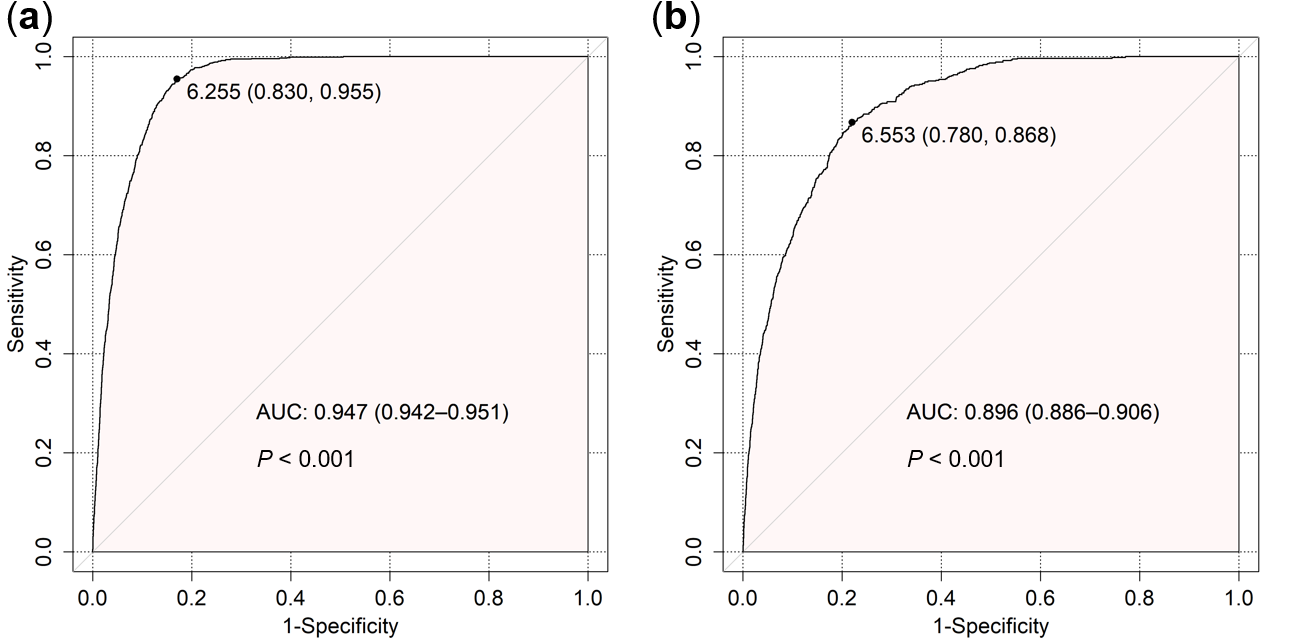

**eFigure 3. Algorithm performance for identifying individuals with CAIDE dementia risk score ≥6 points in the internal validation dataset (a) and the external validation dataset (b).**

Individuals with CAIDE dementia risk score ≥6 points were defined as eligible for multidomain intervention. The points on line indicate the maximum Youden index.

Abbreviation: AUC = area under the receiver operating characteristic curve.

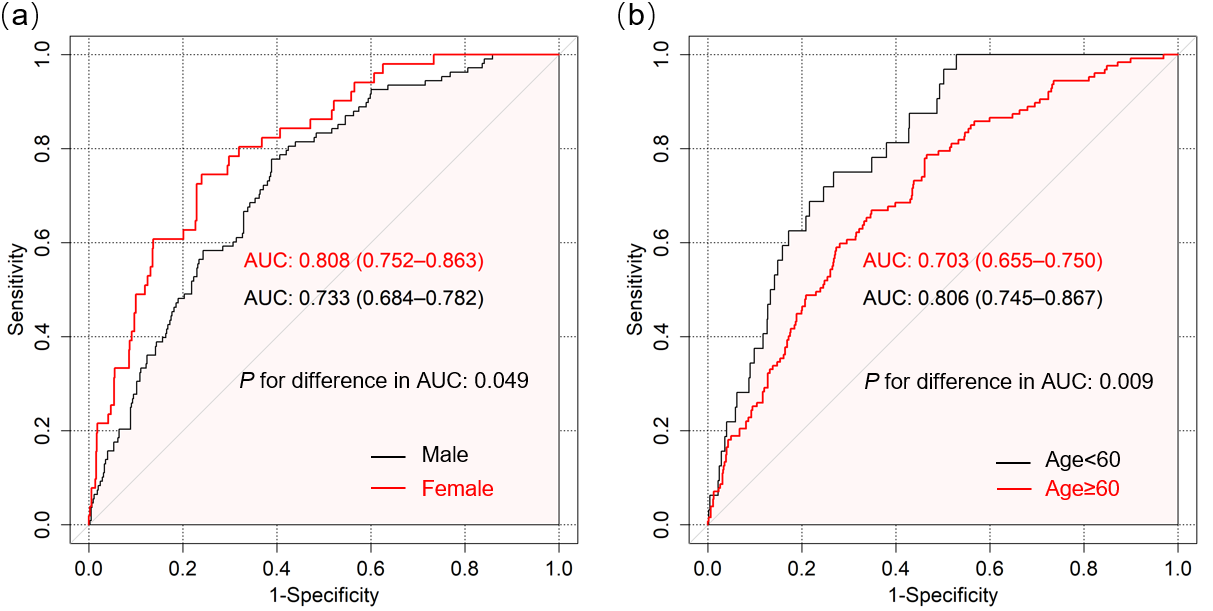

**eFigure 4. Algorithm performance for identifying participants with high dementia risk, subgroup results of sex (a) and age group (b) in the external validation.**

Individuals with high dementia risk were defined as CAIDE dementia risk score ≥10 points. Delong test was used to compare the AUC between subgroups.

Abbreviation: AUC = area under the receiver operating characteristic curve.

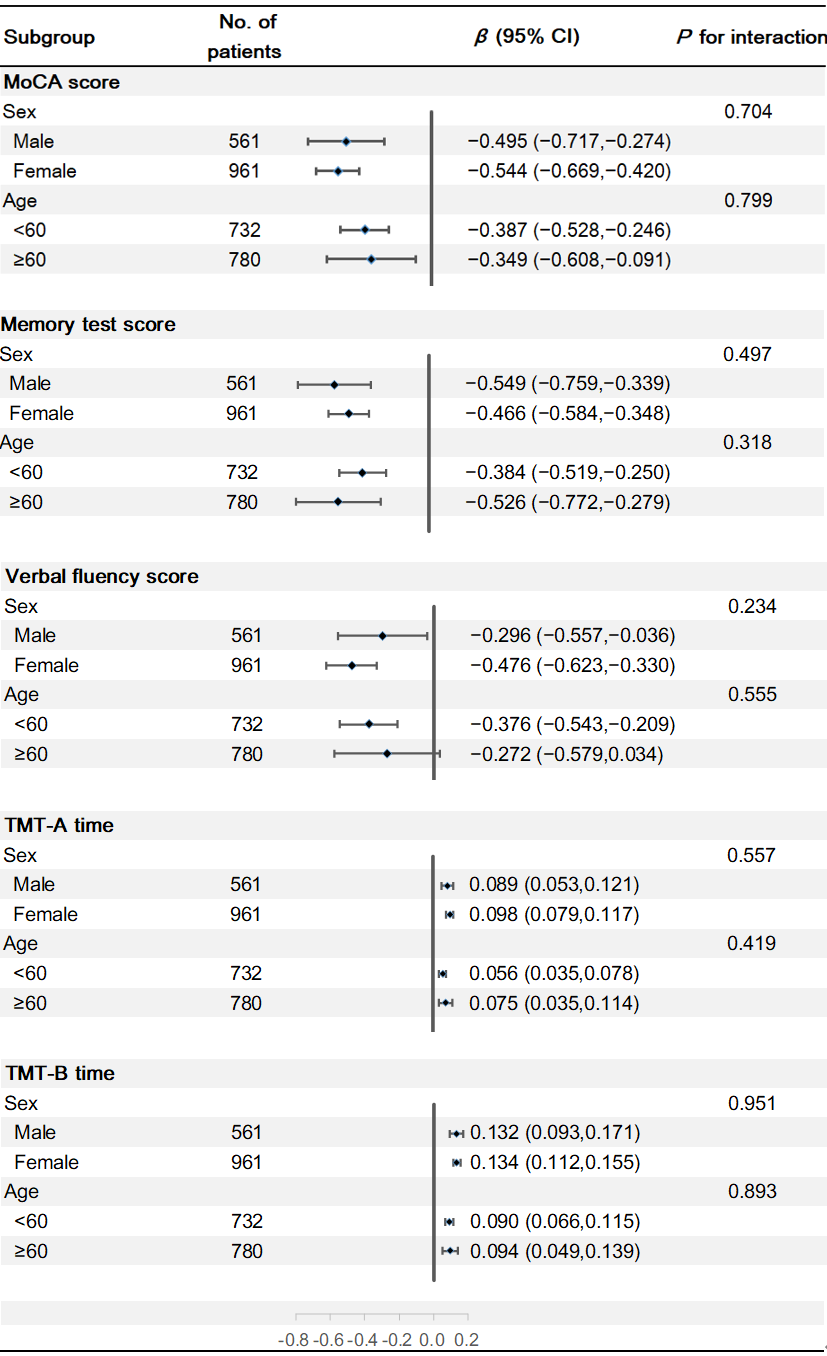

**eFigure 5. Association between estimated CAIDE dementia risk score and different cognitive assessments using multiple linear regression models, subgroup results of sex and age group in the external validation.**

Subgroup analyses were conducted by respectively including the interaction terms of estimated CAIDE dementia risk score with sex, as well as age group in multivariate linear regression models.
